# EEG Biomarkers for ATN Classification in Early Alzheimer’s Disease

**DOI:** 10.64898/2026.06.26.26356640

**Authors:** Claire Braboszcz, Angel David Blanco, Karan Chugani, Victor Fernández, Maitee Rosende-Roca, Laia Cañada, Juan Pablo Tartari, Emilio Alarcón-Martín, Montserrat Alegret, Amanda Cano, Victoría Fernández, Mercè Boada, Xavier Morató, Aureli Soria-Frisch

**Author notes:** Correspondence: Claire Braboszcz < >, Avinguda Tibidabo 47, 08035 Barcelona, Spain.

## Abstract

**INTRODUCTION:** Early detection of Alzheimer’s disease (AD) remains challenging, particularly at preclinical and prodromal stages of subjective cognitive decline (SCD) and mild cognitive impairment (MCI). Electroencephalography (EEG) offers a scalable, non-invasive tool for patient stratification, but its relationship to ATN-defined staging is poorly understood.

**METHODS:** EEG was recorded in 60 participants (SCD, MCI–, MCI+; N=20 per group) using the COGBAT battery (N-back, auditory oddball, 40 Hz auditory steady-state response, resting state). Event-related, spectral, connectivity, and complexity features were compared across groups by ANOVA, correlated with CSF and plasma biomarkers using partial Spearman correlations, and evaluated by univariate ROC and multivariate support vector machine classification.

**RESULTS:** Multiple EEG features discriminated groups and correlated with amyloid and tau biomarkers. P3a amplitude was reduced in MCI+ and tracked CSF *Aβ*, tau, and MMSE. Resting-state theta power was elevated in MCI+, whereas beta power and peak alpha frequency were reduced, the latter correlating specifically with tau. MCI– showed elevated gamma inter-trial coherence, higher signal complexity, and a lower slow-to-fast ratio, consistent with cortical hyperexcitability. Individual features achieved AUCs of 0.72–0.78. EEG added no value for ATN-based discrimination, where plasma pTau217 performed near ceiling (AUC=0.96–0.99), but uniquely separated SCD from MCI (EEG AUC*≈*0.72–0.75) where plasma biomarkers failed (AUC*≈*0.32– 0.33).

**DISCUSSION:** EEG captures ATN-stage-dependent neurophysiological signatures and supports a non-monotonic model of AD progression. It shows promise as a first-pass screening tool at the clinical entry point, where plasma biomarkers are uninformative, identifying individuals who warrant escalation to CSF or PET evaluation.

## 1. BACKGROUND

Alzheimer’s disease (AD) is the leading cause of dementia, accounting for 60–80% of cases [1]. It is a progressive neurodegenerative disorder characterized by a long preclinical phase during which subtle brain changes precede clinical symptoms [2]. Subjective cognitive decline (SCD) and mild cognitive impairment (MCI) represent transitional stages leading to AD dementia, with highly variable clinical trajectories [3]. Individuals with MCI showing amyloid and tau pathology (MCI+) are at higher risk of AD progression, whereas those without such evidence (MCI–) may remain stable [4].

Early detection is essential for timely intervention and clinical trial enrolment, particularly with the advent of disease-modifying therapies [5, 6]. EEG is a scalable, non-invasive technique with documented potential for detecting and characterising AD-related neurophysiological changes across its early stages [7–12].

Current diagnostic approaches for MCI rely on neuropsychological and clinical evaluation, occasionally supported by ATN biomarkers [13]. While informative, these often require invasive and costly procedures, limiting their widespread and repeated use.

Most EEG studies in MCI and AD have focused on resting-state recordings, linking spectral slowing, altered oscillatory dynamics, and signal complexity changes to synaptic dysfunction and cortical disconnection [14–16]. However, resting-state EEG captures only intrinsic brain activity, may miss impairments emerging under cognitive load, and has known limitations in test-retest reliability. EEG classification studies have achieved AUCs of 0.70–0.85 using resting-state features [10–12], with ERP-based approaches showing comparable performance [9, 17]. Yet nearly all such studies stratify groups solely on neuropsychological criteria, without ATN biomarker confirmation, and few directly benchmark EEG against plasma or CSF references.

Task-based EEG probes brain function under cognitive load, providing complementary information to resting-state measures. ERPs show increased latency and reduced amplitude with advancing AD pathology [18]. Here we propose the COGBAT (COG-nitive BATtery), combining resting-state recordings with three task paradigms sensitive to MCI and early AD: an **n-back task** for working memory [19, 20], an **auditory oddball task** for attention and novelty processing [17, 21], and a **40 Hz auditory steady-state response (ASSR)** for neural synchrony and cortical network integrity [22–24].

The relationship between EEG features and fluid biomarkers remains poorly characterised. Resting-state studies have linked theta power to CSF *Aβ* and alpha/beta slowing to tau [25, 26], but ERP-based studies integrating CSF or plasma biomarkers are rare [27]. A recent systematic review underscores that multimodal electrophysiology-biomarker studies remain scarce and that electrophysiological signatures evolve in a non-monotonic, stage-dependent fashion not yet well characterised across ATN-stratified cohorts [28].

Against this background, the present study addresses three key gaps: (i) the absence of ATN-stratified EEG classification studies benchmarking performance against plasma and CSF biomarker references; (ii) the lack of multimodal designs combining resting-state and task-related EEG with fluid biomarkers; and (iii) the unclear complementarity of EEG and plasma biomarkers across distinct clinical questions—distinguishing ATN status within MCI versus separating SCD from MCI irrespective of biomarker profile. We evaluate resting-state and task-related EEG features from the COGBAT battery in individuals with SCD, MCI–, and MCI+, examining associations with fluid biomarkers to assess EEG as a scalable, cost-effective modality for early detection, stratification, and longitudinal tracking of AD-related neurophysiological change.

## 2. METHODS

### 2.1. Participants

All participants were volunteers recruited from patients who underwent cognitive evaluation in the Memory Unit of the Ace Alzheimer Center Barcelona. A total of 60 participants were included and equally distributed across three groups (*N* = 20 each): **Subjective cognitive decline (SCD):** 10 females, mean age 69 *±* 7 years; **MCI with negative A-T-N-biomarkers (MCI–):** 10 females, mean age 72 *±* 6 years and **MCI with positive A+T+(N) biomarkers (MCI+):** 10 females, mean age 75 *±* 5 years.

The ATN classification was based on cerebrospinal fluid (CSF) quantification of *Aβ*1-42, pTau181, and t-Tau. All participants provided written informed consent. The study was approved by the Ethics Committee of the Universitat Autònoma de Barcelona (UAB; reference CEEAH 6252) and by the Ethics Committee of the University Hospital of Bellvitge (CEIm; reference ICPS021/123).

Exclusion criteria included: history of neurological disorders (stroke, epilepsy, meningoencephalitis, brain tumour, severe traumatic brain injury, multiple sclerosis), psychiatric disorders within the past 10 years (bipolar disorder, post-traumatic stress disorder, major depression, psychosis, or suicide attempt), and significant substance use (heavy alcohol consumption, MDMA, amphetamines, cocaine, opioids, benzodiazepines, or cannabis).

### 2.2. Clinical assessment, biomarkers, and group definition

All participants underwent the standard diagnostic procedure of the ACE Alzheimer Center Barcelona. This included an initial neurological and neuropsychological evaluation with the following instruments: Mini-Mental State Examination (MMSE), the memory subtest of the 7-Minute Screen Test [29], the Neuropsychiatric Inventory Questionnaire (NPI; [30]), the Clinical Dementia Rating scale (CDR; [31]), and the Neuropsychological Battery of the ACE Center (NBACE; [32]). The NBACE is a comprehensive battery assessing multiple cognitive domains: attention (WAIS-III digit span forward and backward), automatic inhibition (SKT errors and time), executive functions (category and letter fluency), language (verbal comprehension and visual naming), orientation (global and temporal), praxis (block design WAIS-III, imitation, ideomotor), verbal memory (delayed recall, learning, recognition memory), and visual perception (Luria’s clock and Poppelreuter’s test).

Demographic data (sex, years of education) were obtained from medical records. Participants were classified as MCI if they showed at least one impaired score on the NBACE; those without impairment were classified as SCD.

For all participants, APOE genotype and plasma levels of pTau181 and pTau217 were measured. Participants diagnosed with MCI also underwent anatomical MRI and lumbar puncture to obtain CSF measures of *Aβ* and tau. Based on these results, they were further classified according to the ATN framework [13, 33]. Specifically, participants were assigned to the MCI+ group if their *Aβ*1-42/*Aβ*1-40 ratio was *<* 0.063, and to the MCI– group otherwise.

### 2.3. Experimental protocol

EEG data were acquired using an Enobio-32 device (Neuroelectrics) with gel electrodes placed in the standard 10-20 system, with CMS/DRL reference electrodes placed on the right mastoid and sampling rate 500Hz. The device was connected to the recording computer with a USB cable and data were acquired using the NIC2 software provided by Neuroelectrics. One channel ECG was recorded with sensor placed on the left inner wrist. Participants were seated in a comfortable chair with a laptop computer positioned on the desk directly in front of them. EEG data were acquired continuously while the participant completed a battery of computer-based tasks. The Presentation software (Neurobs, version 21.1 09.05.19) was used to present each task and task instruction. Event triggers were transmitted from the Presentation software to the EEG recording system (NIC2) via the Lab Streaming Layer (LSL) protocol to ensure precise synchronization of stimulus presentation with EEG data acquisition.

### 2.4. COGBAT battery

The COGBAT battery was presented as follows, in the same fixed order for all participants: a 10-min N-back task with 0-and 1-back conditions, an 8-min active three-stimuli auditory oddball task, 5-min of resting state with interleaved eyes opened and eyes closed period (for each, 6 periods of 30 seconds) and a 3-min Auditory Steady State Response (ASSR) task. In between each task, participants could take a self-paced break period. Instructions were given before the start of each specific task, and training was provided before the start of the N-back and auditory oddball task. An experimenter was present at all time near the participants, making sure they understood the instructions. When needed, the responses of the participants were given using a keypad. Participants wore headphones for all tasks involving auditory stimulation or instructions (auditory oddball, resting state, and ASSR).

#### N-back

The N-back task is a continuous working memory paradigm in which participants judged whether the current stimulus matched (target) or differed from (non-target) a stimulus presented N trials earlier. Visual stimuli consisted of single digits (0–9) presented sequentially in white on a black background for 0.6 s, with an inter-stimulus interval of 1.4 s. Responses were made with the right hand using two designated keys: a red key for target stimuli and a blue key for non-target stimuli. In the 0-back condition (low working memory load), a randomly selected digit (e.g., “7”) served as the target throughout the block; participants pressed the target key whenever this digit appeared and the non-target key otherwise. In the 1-back condition, participants indicated whether the current digit was identical to or different from the digit presented immediately prior. The task comprised two consecutive blocks of the 0-back condition followed by two blocks of the 1-back condition, with each block consisting of 50 trials (100 trials per condition in total). Instructions and practice trials were provided prior to the first block of each condition, and participants were permitted a self-paced break at the end of each block.

#### Auditory oddball

In the three-stimulus auditory oddball paradigm, participants were presented with a sequence of 450 auditory stimuli, consisting of frequent tones (1.5 kHz, 200 ms; 80% of trials), target oddball tones (2 kHz, 200 ms; 10% of trials), and novel, non-repeating noises (10% of trials). Stimuli were separated by a fixed inter-stimulus interval of 700 ms. Participants were instructed to respond to target tones by pressing the red key on the keypad as quickly and accurately as possible, while withholding responses to frequent and novel stimuli. Task instructions and practice trials were provided prior to the start of the task.

#### Resting state

Resting-state EEG activity was recorded during a 25-second interleaved eyes-open and eyes-closed paradigm, with each condition repeated six times. Participants received instructions both audibly and visually via text on the screen indicating when to open or close their eyes. During the eyes-open condition, participants were instructed to look at a fixation cross on the screen, whereas during the eyes-closed condition, they were asked to relax and keep their eyes gently closed. No additional task was required during either condition.

#### Auditory Steady-State Response (ASSR)

Auditory steady-state responses were elicited using trains of auditory stimuli. Participants were presented with 160 trials of 40 Hz click trains, each lasting 500 ms, with an inter-trial interval of 750 ms. The stimuli were standard, unattended auditory clicks, and participants were instructed to maintain fixation on a central cross during stimulus presentation.

### 2.5. EEG preprocessing

EEG data from all tasks underwent the same core preprocessing steps for artifact detection and rejection using consistent main parameters, with some task-specific parameters (e.g., additional filtering, epoch length) applied as needed. For each task, data were segmented and processed using a fully automated Python-based pipeline, which included frequency-domain filtering, independent component analysis (ICA) for eye-movement artifact removal, and automated artifact rejection.

Common preprocessing steps across tasks included ICA decomposition and identification of eye-movement–related components and threshold-based automated rejection of artifactual data, defined as any epoch with amplitude exceeding 100 ţV or epochs filtered in the beta band exceeding 30 ţV.

#### General preprocessing

Continuous EEG data were first filtered between 1–45 Hz using a FIR filter. ICA decomposition was performed using the extended Infomax algorithm [34] as implemented in MNE-Python [35]. Independent components (ICs) that showed maximal correlation with frontal channels (FP1 and FP2) were flagged as eye-movement–related and saved for later rejection. The ICA decomposition matrix and indices of ICs to reject were stored for application to each task-specific dataset.

#### Task-specific preprocessing

##### N-back and Auditory Oddball

Data were filtered between 0.05–10 Hz using an IIR filter. Previously computed ICA weights were applied, and eye-movement ICs were rejected. Data were re-referenced to virtual linked mastoids, averaging channels P7, P8, T7, and T8. Epochs were extracted relative to stimulus onset ([-300, 1700] ms for N-back; [-200, 600] ms for auditory oddball) with removal of the pre stimulus baseline period and threshold-based artifact rejection was performed.

#### Resting State

Data were filtered between 1–45 Hz using a FIR filter. ICA weights were applied, and eye-movement ICs rejected. Signals were re-referenced to the common average, and the signal periods corresponding to the eyes-open and eyes-closed conditions were analyzed separately. To minimize artifacts related to the transition between eye states, the first two seconds of each period were excluded. The data were segmented into 4-second epochs with 50% overlap, followed by threshold-based artifact rejection.

#### ASSR

Data were filtered into narrow bands (delta 1–4 Hz, theta 4–8 Hz, alpha 8–12 Hz, beta 12–30 Hz, gamma 35–45 Hz). ICA weights were applied and eye-movement ICs rejected. Epoch trials were extracted relative to stimulus presentation ([-300, 1250] ms) with removal of the pre stimulus baseline period, and threshold-based artifact rejection was applied.

### 2.6. ERP computation

For each task, event-related potentials (ERPs) were computed for each subject and stimulus type, provided that at least 20 trials remained after artifact rejection. The time windows for analysis were defined based on the across-group average of the ERPs of interest. ERP components mean amplitude across respective time windows of interest were computed for analysis.

#### Auditory oddball

Difference waves were computed at electrodes Fz, Cz, and Pz: between Novel and Frequent trials to study the P3a and N2a components, and between Target and Frequent trials to study the P3b and N2b components. The post-stimulus time windows of interest were defined as follows: P3b: 450–600 ms; P3a: 290–530 ms; N2a: 100–300 ms; N2b: 230–350 ms.

#### N-back

Difference waves were computed at central, parietal, and posterior electrodes (C3, Cz, C4, CP1, CP2, P3, Pz, P4) for correctly answered Target and Non-Target trials in both the 0-back and 1-back conditions. Following Mamani et al. [36], the ERP to Non-Target trials (which were more numerous) was used to analyze the P450 component at these same electrodes.

The PNwm component [19] was also computed at these electrodes by subtracting the ERP to all correctly answered trials in the 0-back condition (low memory load) from the ERP to all correctly answered trials in the 1-back condition (high memory load). This difference highlights working memory-related activity by isolating the neural response associated with increased memory load.

### 2.7. Inter-trial coherence

In the ASSR task, inter-trial coherence (ITC) was computed at midline electrodes (Fz, Cz, Pz) from narrow-band filtered trials. The analytic signal was obtained using the Hilbert transform and normalized by its instantaneous amplitude to yield unit phase vectors. These unit phase vectors were averaged across trial epochs, and baseline correction was applied to obtain the ITC, which reflects the consistency of phase across trials at each time point.

For each participant, a single ITC feature value was obtained by identifying the maximum peak dynamic of the ITC across averaged trials at these electrodes.

### 2.8. PSD computation

The PSD was estimated using a Welch-like method, applying a Hamming window to 4-second epochs with 50% overlap and averaging the resulting periodograms across epochs. Then, we extracted the average power within specific frequency bands for each electrode, including delta (1–4 Hz), theta (4–8 Hz), alpha (8–13 Hz), beta (13–30 Hz), and gamma (30–40 Hz) bands.

We averaged these computed features across several electrode clusters (Frontal Cluster: F3, F4, F7, F8, and Fz channels; Central Cluster: Cz, C3, and C4 channels; Temporal Cluster: T7 and T8 channels; Parietal Cluster: P7, P8, P3, P4, and Pz channels; Occipital Cluster: O1 and O2 channels; Midline Cluster: Fz, Cz and Pz channels; Global Cluster: all channels)

Each cluster was further divided into left and right hemispheric subclusters (e.g., left frontal: F3, F7; right frontal: F4, F8). Additionally, electrodes located along the midline were grouped into three separate midline clusters: frontal (Fz), central (Cz), and parietal (Pz). Finally, a global cluster including all electrodes was also computed.

For each cluster and subcluster, we derived the following features: Theta-to-Alpha Ratio (TAR); Slow-to-Fast Ratio (SFR), computed as (*δ* +*θ*)*/*(*α*+*β*); Relative Band Power (RBP), defined as the proportion of power within a specific frequency band relative to the total power across all bands; Peak Alpha Frequency (PAF), identified as the frequency with maximum power within the 5– 15 Hz range (this extended range was chosen to account for potential alpha slowing commonly observed in Alzheimer’s disease); Functional Connectivity Metrics, connectivity between electrode pairs within and across clusters was estimated using phase-locking value (PLV). For each frequency band, connectivity values were averaged across all possible electrode pairs between the corresponding clusters, providing complementary measures of amplitude-and phase-based synchronization; Complexity Metrics: Higuchi Fractal Dimension (HFD) and Waveform Complexity (WC) for each cluster.

### 2.9. Statistics

Group differences between SCD, MCI– and MCI+ were assessed using one-way ANOVA (*p < .*05), unless stated otherwise. Homogeneity of variance was assessed using Levene’s test; when the assumption was violated (*p < .*05), Welch’s ANOVA was used in place of the standard one-way ANOVA. Prior to statistical testing, outliers were identified and removed using the Tukey interquartile range (IQR) method. For significant ANOVAs, post-hoc pairwise comparisons were conducted using Tukey’s honestly significant difference (HSD) test, or Games-Howell test when Welch’s ANOVA was used. For the resting-state analysis, which surveyed all combinations of electrode cluster, condition and frequency band without regional hypotheses, ANOVAs were performed separately for each cluster, condition (eyes open, eyes closed) and frequency band, and *p*-values were adjusted using the false discovery rate (FDR) procedure [37] to account for multiple comparisons. Task-related ERP and ASSR analyses targeted components and electrodes selected on the basis of prior literature and are reported uncorrected.

We also examined associations between the extracted EEG features from each task and selected clinical or biomarker variables using partial Spearman correlations, with age included as a covariate [38]. Outliers were removed with the IQR Tukey rule, and *p*-values were adjusted for multiple comparisons using the FDR procedure, unless stated otherwise. The analyses included all CSF and plasma biomarkers as well as the MMSE clinical score.

Significant associations were visualized using heatmaps, where the cell color represents the correlation coefficient (*r_s_*).

### 2.10. Classification Analysis

#### 2.10.1. Univariate ROC Analysis

To evaluate the discriminative ability of individual EEG and clinical features, receiver operating characteristic (ROC) curves and corresponding area under the curve (AUC) values were computed for each feature separately [39]. Three binary classification contrasts were examined: MCI– versus MCI+; SCD and MCI– combined versus MCI+; and SCD versus MCI– and MCI+ combined. Features were drawn from the five domains described above: auditory oddball ERP components, N-back task ERP amplitudes and performance measures, ASSR metrics (evoked power and ITC in the low-gamma band), resting-state EEG markers, and clinical and fluid biomarker variables. Features were included regardless of whether they reached statistical significance in the group-level ANOVA. Missing values were excluded prior to analysis on a per-feature basis. For a subset of features whose raw AUC fell below 0.50 — indicating that higher feature values were paradoxically associated with the reference class — group coding was reversed prior to computing the ROC curve, so that AUC values consistently reflect discriminative capacity irrespective of feature polarity.

#### 2.10.2. Multivariate SVM Classification

To assess the added value of combining features across modalities, a support vector machine (SVM) classifier with a radial basis function (RBF) kernel was applied across three binary classification contrasts: (1) SCD versus MCI (MCI– and MCI+ combined); (2) SCD and MCI– combined versus MCI+; and (3) MCI– versus MCI+. For the third contrast, the CSF *Aβ*1-42/*Aβ*1-40 ratio was additionally included as a candidate feature given its direct relevance to amyloid pathology staging.

For each contrast, three multimodal feature set combinations were evaluated: (i) best-performing EEG features combined with behavioural measures; (ii) plasma pTau217 combined with best EEG features; and (iii) plasma pTau217 combined with best EEG features and behavioural measures. The best-performing EEG features were selected based on the AUC values obtained in the univariate ROC analysis described above, retaining the single EEG feature with the highest individual discriminative ability for the corresponding contrast. Prior to classification, all features were standardised using z-score normalisation. Features were verified to have non-missing values across all participants within each group before inclusion; any model for which no valid feature set could be constructed was excluded from that contrast.

Classification performance was evaluated using 100-iteration stratified hold-out cross-validation. Across iterations, ROC curves were computed and averaged to yield a mean ROC curve with a corresponding standard deviation band, and AUC values were recorded per iteration. Mean AUC and its standard deviation across the 100 iterations are reported for each model and contrast.

#### 2.10.3. Model Comparison

To contextualise multimodal classification performance, AUC distributions from the 100-iteration hold-out procedure were summarised as boxplots for each contrast separately, with models ordered by descending mean AUC. Horizontal reference lines indicate single-feature AUC values derived from the univariate ROC analysis described above. For all three contrasts, reference lines are shown for plasma pTau217, plasma pTau181, and the best-performing individual EEG feature. Additionally, MMSE is included as a reference for the SCD versus MCI contrast, given its clinical relevance as a screening measure in this comparison. For the MCI– versus MCI+ contrast, the *Aβ*1-42/*Aβ*1-40 ratio is additionally shown as a reference, reflecting its direct relevance to amyloid pathology staging in this subgroup. This allows direct visual assessment of whether combining EEG, behavioural, and fluid biomarker features provides added discriminative value beyond any single modality alone.

## 3. RESULTS

### 3.1. ERP results

#### 3.1.1. Auditory oddball

In the behavioural performance during the auditory oddball task (see Table S1, Supplementary Material), significant group effects were observed for accuracy (Welch’s *F* (2, 28.80)= 9.37, *p* < 0.001, 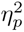 = 0.32) but not response time (*F* (2, 52)= 0.02, *p* = 0.983). Mean accuracy in the MCI+ group was significantly lower than in the MCI– (*p* = 0.010) and SCD groups (*p* = 0.001). There was no significant difference between the MCI– and SCD groups (*p* = 0.19).

Table 1 reports the ERP and ASSR feature statistics for all tasks. Significant group differences were observed in the mean peak amplitude of the P3a component at electrode Cz in the 290–530 ms time window (*F* (2, 55)= 4.90, *p* = 0.011, 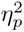 = 0.15) (Figure 1 A and B). The MCI+ group exhibited smaller P3a amplitudes compared to both MCI– (*p*= 0.023) and SCD (*p*= 0.021), whereas no difference was observed between SCD and MCI–. No significant group differences were found at electrode Pz (*F* (2, 53) = 2.86, *p*= 0.06). No significant group differences were observed for the P3b component (450–600 ms) on Pz, nor for the N2a (Novel–Frequent, 100–300 ms) or N2b (Target–Frequent, 230–350 ms) components at Cz or Fz; the P3b and N2b waveforms and amplitude distributions are shown in Figure S1 (Supplementary Material). We did not observe significant group difference in the latency of any of the analyzed components.

**Figure 1.**
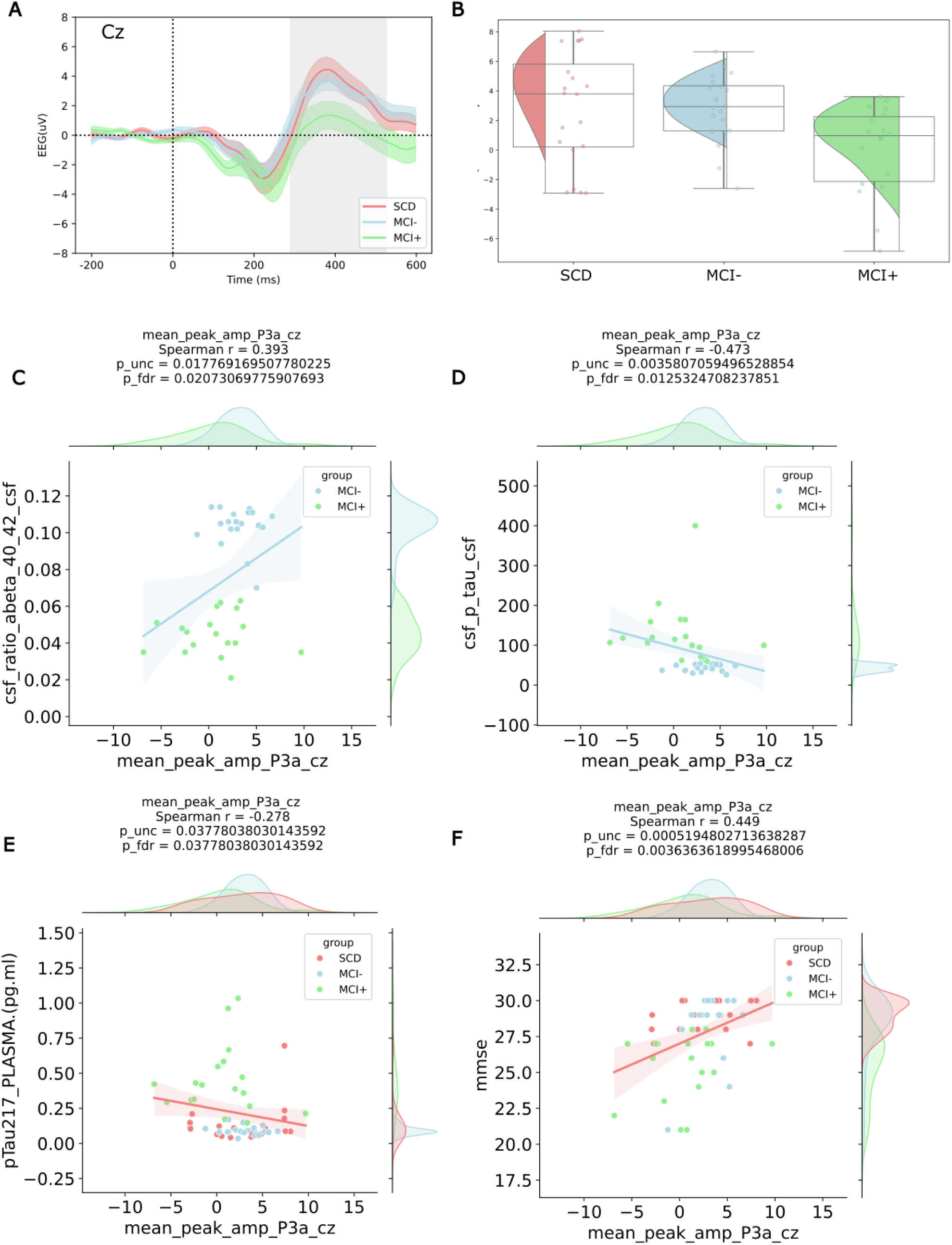
Auditory Oddball — P3a ERP on Cz. **A.** P3a ERP on Cz for each group. The 290–530 ms time window of interest is shown as a grey area. **B.** Mean peak amplitude distribution of P3a on Cz for each group over the time window of interest. The MCI+ group shows significantly lower mean amplitude than the SCD and MCI*−* groups. **C–F.** Significant correlation between the P3a mean amplitude on Cz and CSF *Aβ*40*/*42 ratio (panel C), CSF ptau (panel D), plasma ptau 217 (panel E) and MMSE (panel F).

**Table 1.**
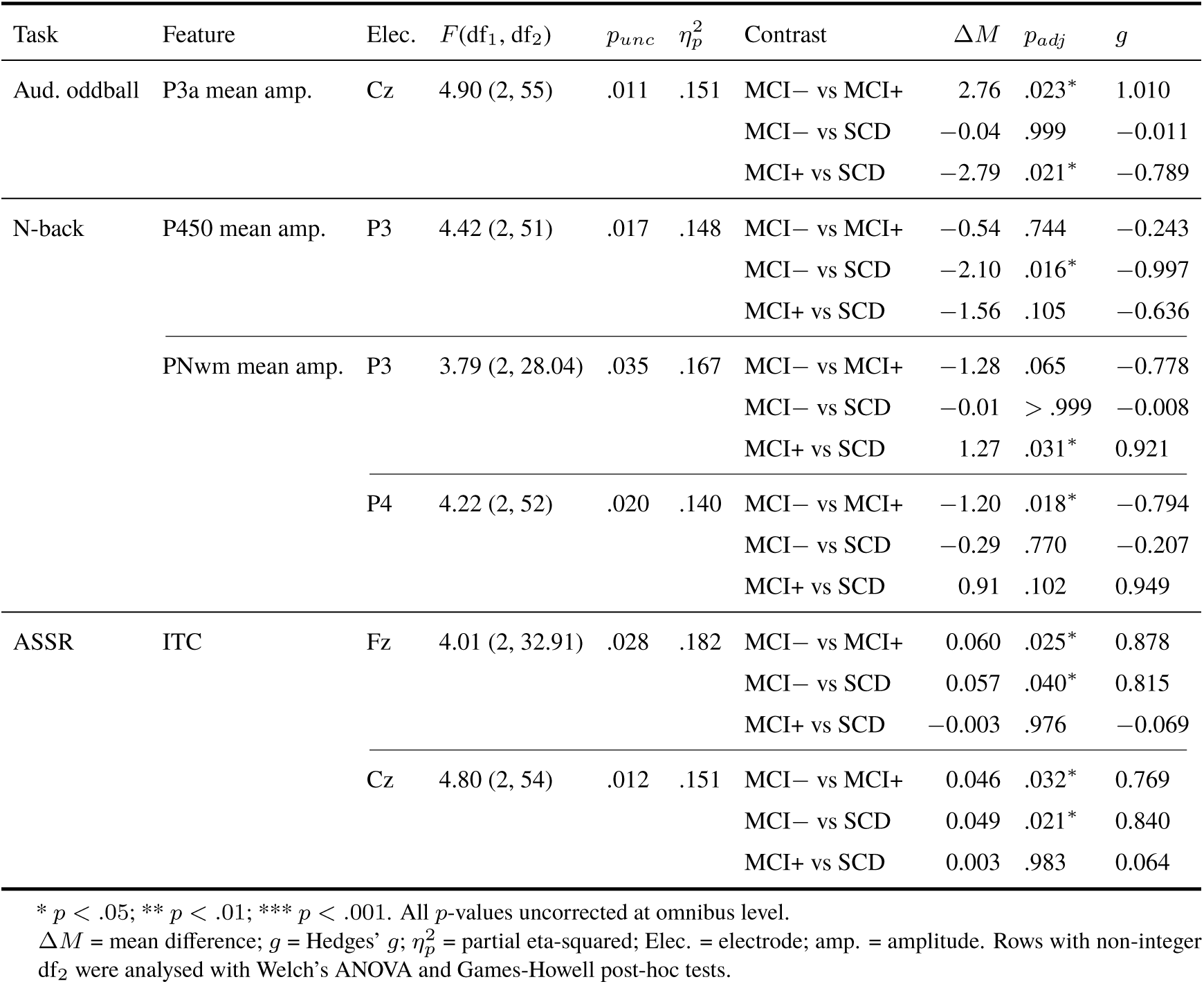
ERP and EEG feature ANOVA results and post-hoc tests. (Tukey HSD, or Games-Howell where Welch’s ANOVA was used). Effect sizes: 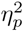 (omnibus), Hedges’ *g* (contrasts).

#### 3.1.2. N-back

A mixed-design ANOVA was conducted on the accuracy to the N-back task with Group (SCD, MCI–, MCI+) as a between-subjects factor and Condition (0-back, 1-back) as a within-subjects factor (see Table S1, Supplementary Material). There was a significant main effect of Group, *F* (2, 48)= 13.39, *p*< 0.001, 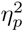 = 0.36, indicating overall differences in error rates between groups. There was also a significant main effect of Condition, *F* (1, 48)= 59.56, *p* < .001, 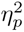 = 0.55, showing that participants made more errors in the 1-back condition compared to the 0-back condition. Importantly, the Group *×* Condition interaction was significant, *F* (2, 48)= 6.28, *p*= 0.004, 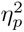 = 0.21, indicating that the effect of condition differed across groups.

Follow-up pairwise comparisons revealed that in the 0-back condition, the MCI+ group made more errors than the SCD group (*p*=0.01), whereas the MCI+ and MCI– groups did not differ significantly (*p*=0.06). In the 1-back condition, the MCI+ group made more errors than both the SCD (*p*=0.001) and MCI– (*p*=0.004) groups.

A 3 (Group) *×* 2 (Condition) mixed ANOVA was conducted on reaction time with Group as the between-subjects factor and Condition as the within-subjects factor. There was a significant main effect of Group, *F* (2, 51)= 13.44, *p*< 0.001,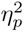 = 0.34, indicating that mean reaction times differed significantly across the three groups. There was also a significant main effect of Condition, *F* (1, 51) = 49.27, *p* < .001, 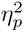 = 0.49, indicating that reaction times differed significantly between the two conditions. Importantly, the Group *×* Condition interaction was significant, *F* (2, 51) = 3.23, *p* = .048, 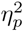 = 0.11.

In both the 0-back and 1-back condition the MCI+ group is slower to respond than the SCD (*p*<0.005 in both conditions) and the MCI– groups (*p*<0.005 in both conditions). There was no difference in response time between SCD and MCI– group in any condition (*p*=0.97 in 0-back and *p*=0.59 in 1-back condition).

Significant group differences were observed in the P450 component during Non-Target correct trials in the 1-back condition (*F* (2, 51)=4.42, *p*=0.017, 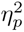 = 0.15; full ERP statistics in Table 1) . At electrode P3, the MCI– group showed reduced P450 amplitude compared to the SCD group in the 350–720 ms time window (*p* < 0.05). Although the MCI+ group did not differ significantly from the SCD group, its mean amplitude distribution was more similar to that of the MCI– group. No significant differences were found in the 0-back condition or at other electrode sites.

The PNwm component, computed as the difference wave between all correct trials in the 1-back (high working memory load) and 0-back (low working memory load) conditions, was absent in the MCI+ group but present in the MCI– and SCD groups, replicating the findings of Missonnier et al. [19]. Significant PNwm differences were observed in the 320–597 ms time window at electrodes P3 (Welch’s *F* (2, 28.04)=3.79, *p*=0.035, 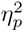=0.17) with amplitude of the negative peak in MCI+ group significantly reduced compared to SCD (*p*=0.03) and at electrode P4 (*F* (2, 52) = 4.22, *p*=0.020, 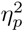 =0.14) with amplitude of the negative peak reduced in MCI+ compared to MCI– (*p*=0.018), see Figure S2 (Supplementary Material).

### 3.2. ASSR

Significant group differences in the ITC maximum peak dynamic were observed at electrodes Fz (Welch’s *F* (2, 32.91)= 4.01, *p*=0.028, 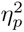=0.18; full ITC statistics in Table 1) and Cz (*F* (2, 54)=4.8, *p*=0.01, 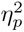 =0.15) (Figure 2). At Fz, the MCI– group exhibited higher maximum peak ITC values than both the SCD (*p* = 0.040) and MCI+ groups (*p* = 0.025). At Cz, the MCI– group also showed higher maximum peak ITC values compared to the SCD (*p*=0.02) and MCI+ groups (*p*=0.03). No significant differences were found at other electrodes.

**Figure 2.**
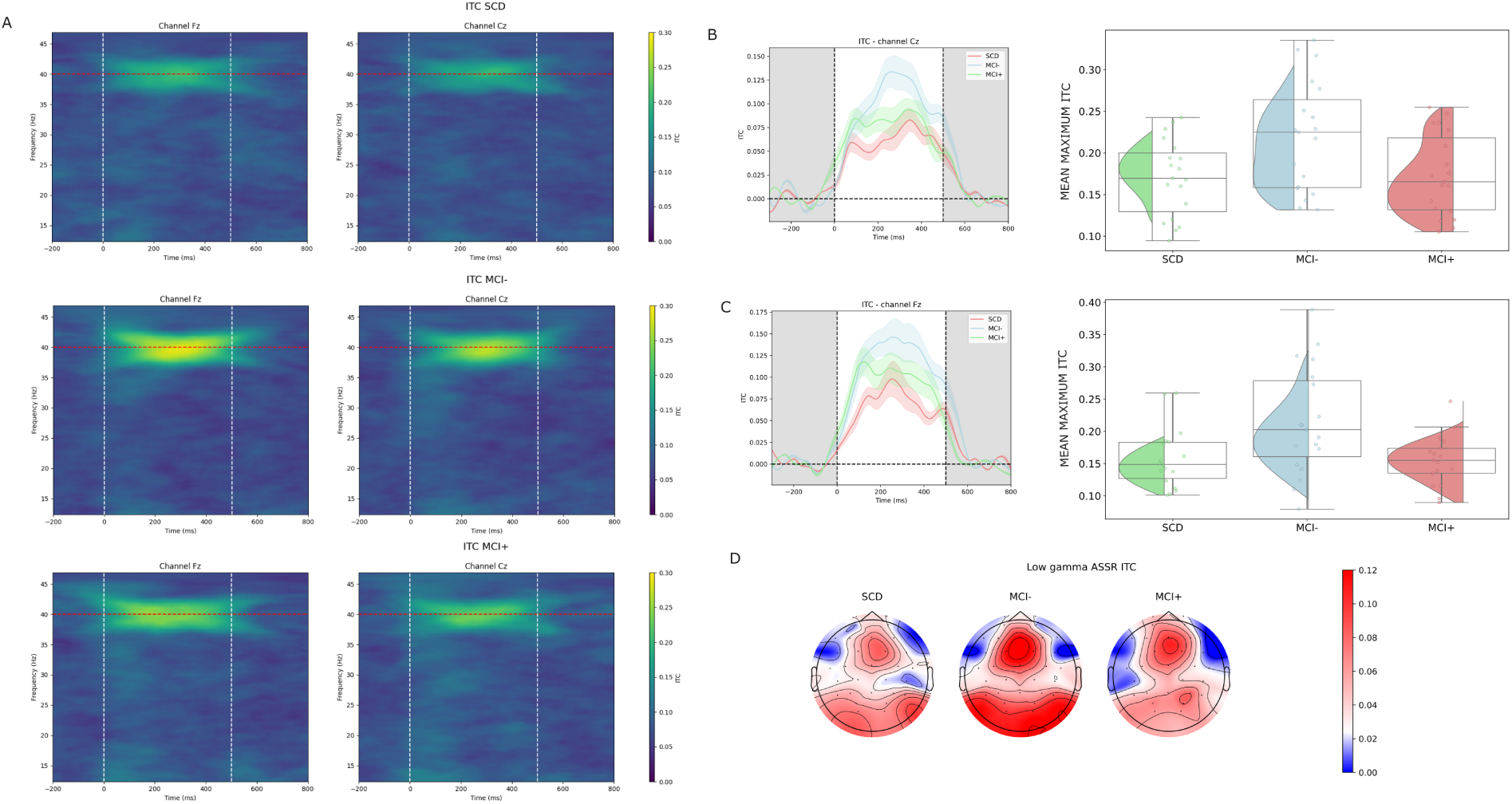
ASSR — ITC. **A.** ITC spectrograms on channels Fz and Cz for each group. The 40 Hz frequency corresponding to stimulation is shown as a red dotted line. Horizontal white dotted lines indicate stimulation duration. **B.** ITC timeplot (right) and distribution (left) per group on channel Cz. ITC in MCI*−* was significantly higher compared to SCD (*p*=0.02) and MCI+ (*p*=0.03) groups. **C.** ITC timeplot (right) and distribution (left) per group on channel Fz. ITC in MCI*−* was significantly higher compared to SCD (*p*=0.04) and MCI+ (*p*=0.02) groups. **D.** Topographical maps of ITC activity per group.

### 3.3. Resting state

#### 3.3.1. EEG spectral activity

Unless specified otherwise, results are reported for the eyes-open condition. Relative band-power topographies and group power spectra are shown in Figure 3A and Figure 3B, respectively. Only effects surviving FDR correction are summarized here, whereas full ANOVA and post-hoc statistics are provided in Table S2 (Supplementary Material).

**Figure 3.**
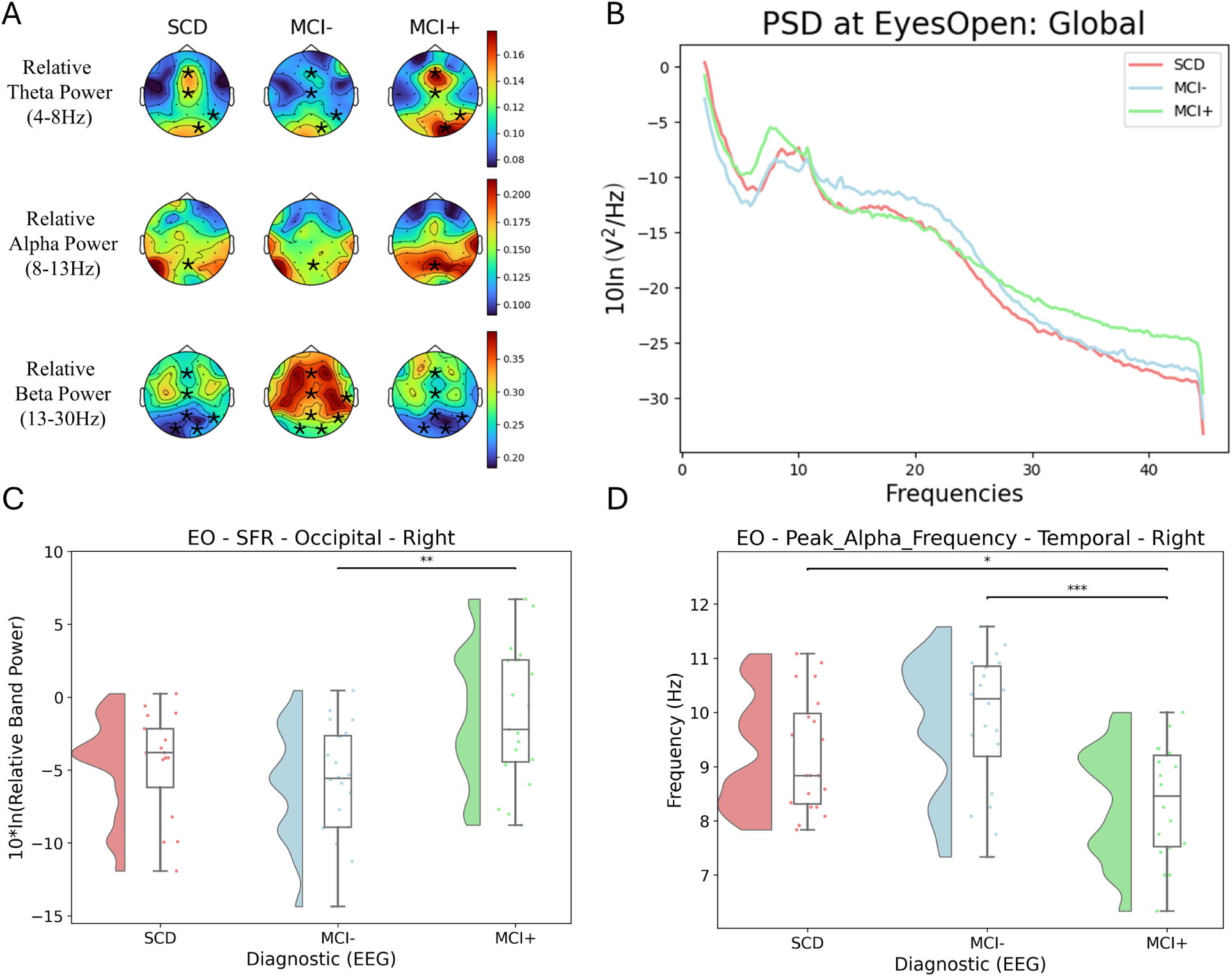
Resting-state EEG results in the eyes-open condition. **A.** Topographical maps of relative theta, alpha, and beta power for SCD, MCI*−*, and MCI+ groups. Areas marked with asterisks indicate regions with significant group differences according to ANOVA. Increased theta and alpha power can be observed in MCI+ compared to the other groups, whereas beta power is higher in MCI*−*. **B.** Power spectral densities (PSD) for each group: SCD in red, MCI*−* in blue, and MCI+ in green. **C.** Barplot of the Slow-to-Fast Ratio (SFR) at right occipital regions, showing significantly higher SFR in MCI+ compared to MCI*−*. **D.** Barplot of peak alpha frequency (PAF) at right temporal regions, indicating lower PAF in MCI+ relative to MCI*−* and SCD. Asterisks indicate significance levels: * *p <*0.05, ** *p <*0.01, *** *p <*0.001.

##### Theta power

Significant group effects were observed in the right parietal (F(2,53)=5.77, p*_FDR_*=0.04, 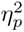 =0.18), right occipital (F(2,55)=5.98, p*_FDR_*=0.04, 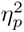 =0.18), midline frontal (F(2,52)=5.32, p*_FDR_*=0.04, 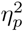 =0.17), and midline central regions (F(2,53)=6.40, p*_FDR_*=0.04, 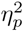 =0.19). Post-hoc analyses showed higher theta power in MCI+ than in MCI– across all significant regions. In addition, MCI+ showed higher theta power than SCD in the right parietal and right occipital regions, while SCD showed higher theta power than MCI– in the midline central region (Table S2).

##### Alpha power

A significant group effect was found in the midline parietal region (F(2,52)=5.42, p*_FDR_*=0.04, 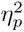 =0.17). Post-hoc comparisons indicated higher alpha power in MCI+ than in both MCI– and SCD, whereas MCI– and SCD did not differ significantly (Table S2).

##### Beta power

Significant group differences emerged in the right temporal (F(2,55)=6.20, p*_FDR_*=0.04, 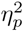 =0.18), right parietal (F(2,55)=6.06, p*_FDR_*=0.04, 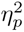 =0.18), left occipital (F(2,55)=5.87, p*_FDR_*=0.04, 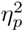 =0.18), right occipital (F(2,56)=7.40, p*_FDR_*=0.04, 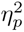 =0.21), midline frontal (F(2,53)=13.11, p*_FDR_*=0.002, 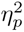 =0.33), midline central (F(2,55)=7.49, p*_FDR_*=0.04, 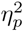 =0.21), and midline parietal regions (F(2,52)=5.30, p*_FDR_*=0.04, 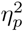=0.17). Post-hoc comparisons consistently indicated reduced beta power in MCI+ compared to MCI– across all significant regions. In posterior and midline regions, SCD also showed reduced beta power relative to MCI–, specifically in the left occipital, right occipital, midline frontal, midline central, and midline parietal regions. The largest effect size was observed in the midline frontal region (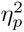 =0.33; Table S2).

##### Slow-to-Fast Ratio (SFR)

Significant effects were detected in the right occipital (F(2,53)=6.25, p*_FDR_*=0.04, 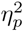 =0.19), midline frontal (F(2,55)=6.94, p*_FDR_*=0.04, 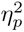 =0.20), and midline central regions (F(2,54)=5.59, p*_FDR_*=0.04, 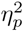 =0.17). Post-hoc analyses showed higher SFR in MCI+ compared to MCI– across all significant regions. Additionally, SCD exhibited higher SFR than MCI– in the midline frontal and midline central regions, whereas in the right occipital region only MCI+ differed significantly from MCI– (Figure 3C; Table S2).

##### Peak alpha frequency (PAF)

Significant group differences in PAF were detected in both resting-state conditions (Figure 3D; full statistics in Table S3, Supplementary Material). In the eyes-open condition, a significant effect emerged in the right temporal region (F(2,55)=5.36, p*_FDR_*=0.049, 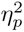 =0.16), driven by lower PAF in MCI+ than in both MCI– (p*_adj_*<0.001) and SCD (p*_adj_*=0.039), with no difference between MCI– and SCD. In the eyes-closed condition, significant effects were found in left frontal, bilateral temporal, bilateral parietal, right occipital, and left central regions. Post-hoc comparisons showed lower PAF in MCI+ than in MCI– in left frontal, bilateral temporal, bilateral parietal, and left central regions, whereas MCI+ and SCD did not differ significantly in any region. MCI– showed higher PAF than SCD in bilateral parietal and right occipital regions.

#### 3.3.2. EEG functional connectivity

Results are reported for the eyes-closed condition, with significant phase-locking value (PLV) differences mapped in Figure 4A. Only effects surviving FDR correction are described below, whereas full ANOVA and post-hoc statistics are provided in Table S4 (Supplementary Material).

**Figure 4.**
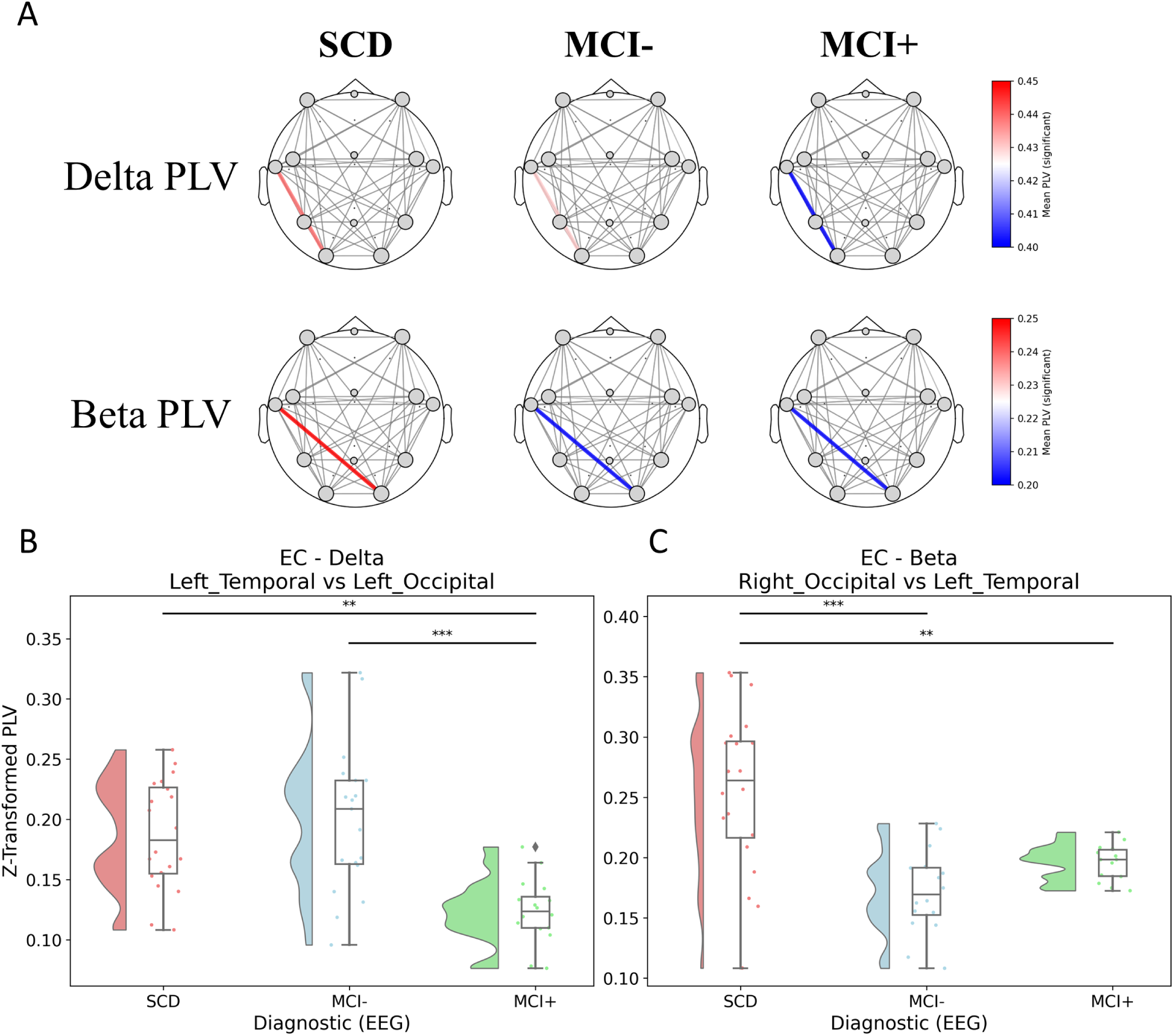
Resting-state EEG functional connectivity in the eyes-closed condition. **A.** Topographical maps of significant Phase-Locking Value (PLV) differences between brain regions for delta and beta bands across SCD, MCI*−*, and MCI+ groups, as identified by ANOVA. **B.** Barplot showing delta-band connectivity between left temporal and left occipital regions, indicating reduced connectivity in MCI+ compared to MCI*−* and SCD. **C.** Barplot showing beta-band connectivity between right occipital and left temporal regions, revealing reduced connectivity in MCI+ and MCI*−* relative to SCD. In both barplots, SCD is shown in red, MCI*−* in blue, and MCI+ in green. Asterisks indicate significance levels: * *p <*0.05, ** *p <*0.01, *** *p <*0.001.

##### Delta connectivity

A significant group effect was observed for delta-band connectivity between the left temporal and left occipital regions (F(2,54)=9.99, p*_FDR_*=0.044, 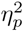 =0.27). Post-hoc analyses revealed reduced delta connectivity in MCI+ compared to both MCI– (Δ*M* =0.067, p*_adj_*<0.001, g=1.305) and SCD (Δ*M* =0.055, p*_adj_*=0.003, g=1.311), whereas MCI– and SCD did not differ significantly (Figure 4B).

##### Beta connectivity

A significant group effect emerged for beta-band connectivity between the right occipital and left temporal regions (F(2,49)=12.70, p*_FDR_*=0.016, 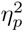 =0.34). Post-hoc comparisons indicated higher connectivity in SCD than in both MCI+ (Δ*M* =0.063, p*_adj_*=0.002, g=1.182) and MCI– (Δ*M* =0.077, p*_adj_*<0.001, g=1.312), whereas MCI+ and MCI– did not differ significantly (Figure 4C). This beta-band connection showed the largest effect size among the significant connectivity findings (2 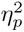 = 0.34; Table S4, Supplementary Material).

#### 3.3.3. Complexity features

##### Higuchi Complexity

During eyes-opened rest, significant group effects were observed in the frontal midline region (F(2,54)=6.92, p*_FDR_*=0.027, 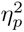 =0.20). Post-hoc analyses indicated that MCI– showed higher Higuchi complexity compared to SCD in frontal midline (p=0.0014). Other contrasts were not significant (MCI+ vs MCI–, p=0.134; MCI+ vs SCD, p=0.191).

##### Waveform Complexity

During eyes-open rest, significant group effects were observed in the left occipital (F(2,54)=6.41, p*_FDR_*=0.021, 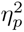 =0.19), right occipital (F(2,53)=5.82, p*_FDR_*=0.023, 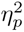 =0.18), and parietal midline regions (F(2,54)=7.59, p*_FDR_*=0.016, 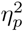 =0.22).

Post-hoc analyses indicated that MCI– showed higher Waveform Complexity compared to SCD in left occipital (p=0.0022), right occipital (p=0.0036), and parietal midline regions (p=0.0008). Other contrasts were not significant (MCI+ vs MCI–, left occipital p=0.283; MCI+ vs SCD, left occipital p=0.113; MCI+ vs MCI–, right occipital p=0.118; MCI+ vs SCD, right occipital p=0.386; MCI+ vs MCI–, parietal midline p=0.178; MCI+ vs SCD, parietal midline p=0.102). These patterns are illustrated in Figure S3 (Supplementary Material), and full statistics are provided in Table S5 (Supplementary Material).

### 3.4. EEG features correlation with clinical scores and biomarkers

Partial Spearman correlations (controlling for age) revealed that several EEG features correlates with clinical and biomarker measures (Figure S4 (Supplementary Material)). After correcting for multiple comparisons, larger auditory oddball P300 ERPs were linked to better cognition and a healthier biomarker profile (see Figure 1 panels C-F), whereas slower EEG rhythms were associated with poorer cognition and higher pathology. Specifically, greater P3a amplitude at Cz (and, to a lesser extent, P3b at Pz) correlated with higher MMSE scores (*r ≈ .*32–.45), higher CSF *Aβ*42 and *Aβ*40*/*42 ratio (*r ≈ .*39–.42), and lower CSF tau, p-tau, and plasma p-tau181 (*r ≈ −.*33 to *−.*47). Right-parietal theta power during eyes-open rest (*RP_Theta_EO*) showed the opposite pattern: higher theta power was linked to lower MMSE and reduced amyloid (*Aβ*42 and *Aβ*40*/*42 ratio; *r ≈ −.*35 to *−.*56), but elevated CSF and plasma tau (*r ≈* +.37 to +.55). Eyes-open beta power across multiple sites (*LO/RO/RP/RT_Beta_EO*) correlated positively with amyloid (*r ≈* +.39 to +.51) and negatively with tau measures (*r ≈ −.*29 to *−.*55). Peak alpha frequency (*RT_PAF_EO* and *LT/LC/LF_PAF_EC*) also tracked lower tau levels (*r ≈ −.*43 to *−.*58).

Indices of slow-to-fast ratio (SFR; *MF/MC/RO_SFR_EO*) were associated with higher tau (*r ≈* +.31 to +.60), lower amyloid (*r ≈ −.*40 to *−.*50), and lower MMSE (*r ≈ −.*36 to *−.*39). Similarly, Higuchi complexity at MF (*MF_Higuchi_EO*) decreased with higher tau/p-tau (*r ≈ −.*46 to *−.*47).

At the uncorrected *p*-value level only (*p*_unc_ *< .*05), auditory oddball larger N2 at Cz were related to lower tau/p-tau (*r ≈ −.*28 to *−.*41), and reduced PNwm amplitudes were linked to lower CSF *Aβ*40*/*42 ratio or lower CSF *Aβ*42 (*r ≈ −.*35 to *−.*44).

### 3.5. Classification models comparison

Figure 5 summarises the AUC distributions across the 100-iteration hold-out procedure for each contrast and model combination.

**Figure 5.**
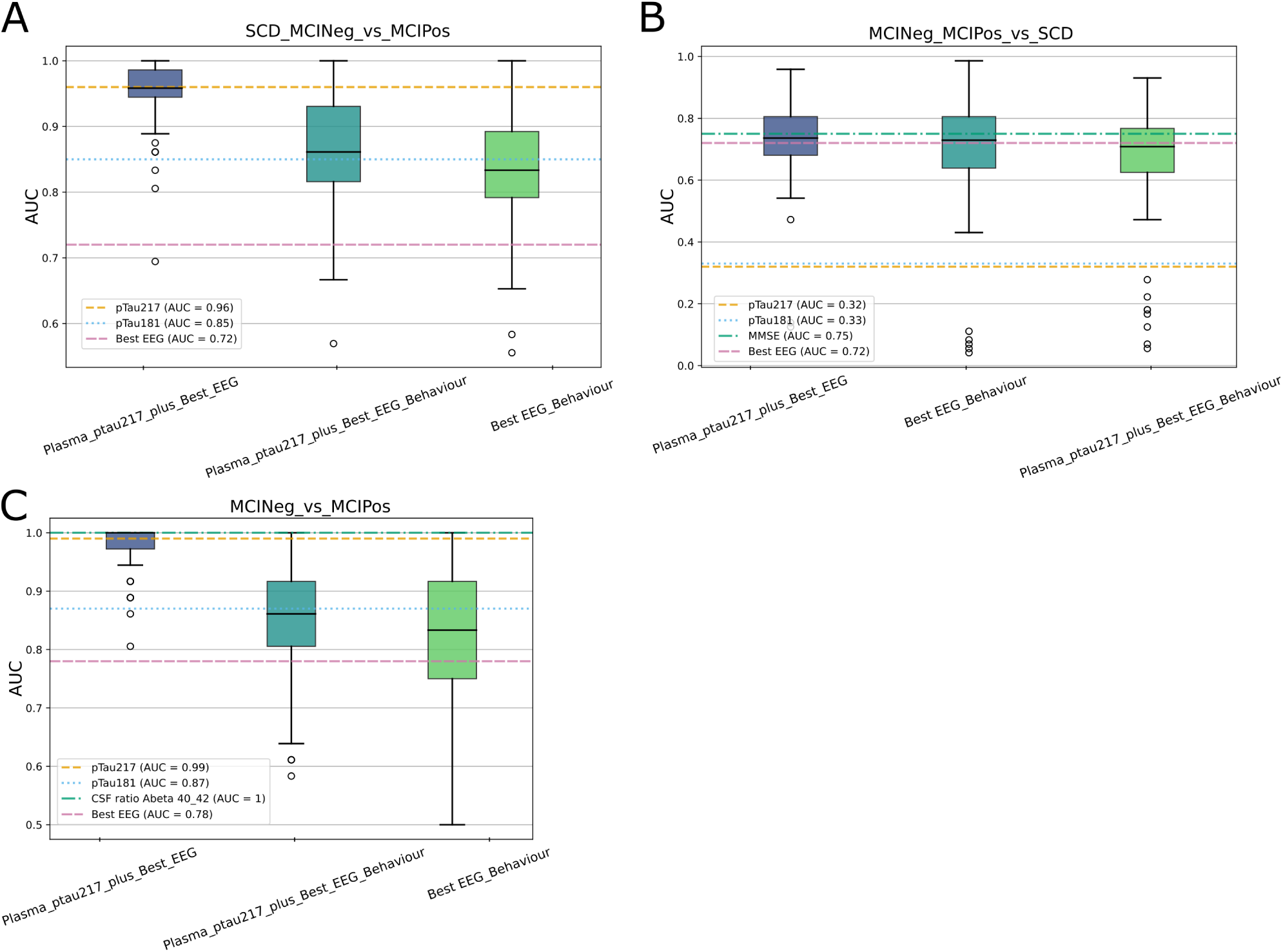
Multivariate SVM classification models comparison. **A.** [SCD and MCI ATN*−*] versus MCI A+T+N+ contrast. Best EEG feature is the right occipital theta power in eyes-open resting state. **B.** [MCI*−* and MCI+] versus SCD contrast. Best EEG feature is the resting-state eyes-closed PLV in the beta band between right occipital and left temporal areas. **C.** MCI*−* versus MCI+ contrast. Best EEG feature is the slow-to-fast ratio measured in the frontal midline during resting-state eyes-open.

For the [SCD MCI and ATN–] versus MCI A+T+N+ contrast (Figure 5A), the best EEG feature is the right occipital theta power in eyes opened rest condition (AUC = 0.72) The model combining plasma pTau217 with this best EEG feature achieved the highest median AUC (median = 0.96), matching the univariate pTau217 reference (AUC = 0.96). Adding behavioural measures to this combination did not improve performance, while the EEG-plus-behaviour model alone yielded a lower median AUC (AUC = 0.83), comparable to the univariate pTau181 reference (AUC = 0.85).

For the [MCI– and MCI+] versus SCD contrast (Figure 5B), the best EEG feature is the resting state eyes closed phase locking value (PLV) in beta band between the right occipital and left temporal areas (AUC = 0.72). All three multimodal models substantially outperformed the univariate plasma biomarker references (pTau217: AUC = 0.32; pTau181: AUC = 0.33), with median AUCs clustering around 0.70–0.74. Performance was broadly comparable across the three models and approached the MMSE reference (AUC = 0.75) and the best individual EEG feature (AUC = 0.72), suggesting that multimodal combinations did not yield substantial gains over these single-modality benchmarks for this contrast.

For the MCI– versus MCI+ contrast (Figure 5C), the best EEG feature is the measure of slow-to-fast ratio in the frontal midline during eyes opened resting state condition (AUC = 0.78). For this contrast, the plasma pTau217 plus best EEG model achieved a near-perfect median AUC (AUC= 1), consistent with the univariate pTau217 (AUC = 0.99) and CSF *Aβ*40*/*42 ratio (AUC = 1.0) references. The two remaining models showed lower and more variable performance (median AUC = 0.86 and AUC = 0.83 respectively), though both remained above the best individual EEG reference (AUC = 0.78).

## 4. DISCUSSION

We investigated the relationship between EEG features, cognitive performance, and AD biomarkers in prodromal AD participants classified as SCD, MCI–, and MCI+. Our results show that specific EEG markers can both distinguish between groups and correlate with amyloid and tau pathology, highlighting their potential as low-cost, non-invasive proxies for prodromal AD biomarker status. The present work contributes to the still sparse body of multimodal studies integrating EEG with fluid biomarkers — a gap recently highlighted by Sohrabpour et al. [28], who underscored that such designs are uniquely positioned to link electrophysiological features to the underlying proteinopathy of AD.

### 4.1. Task-related EEG markers reflect cognitive health and amyloid/tau status

Auditory oddball ERPs, particularly P3a at Cz, were larger in participants with better cognitive scores and healthier biomarker profiles, with the P3a amplitude significantly smaller in MCI+ compared to MCI– and SCD. These findings align with previous literature indicating that P3 amplitude decreases with cognitive decline and AD pathology [18, 40], and our correlations extend this by showing that P3 components track not only cognition (MMSE) but also CSF and plasma *Aβ* and tau.

We did not find significant group differences in the N2a or N2b components. This null result likely reflects the sensitivity of our active three-stimulus paradigm and the early disease stage of our sample; N2 effects may be more prominent in later-stage AD or passive paradigms [17, 21]. The N2a/N2b components may also be less directly tied to amyloid or tau burden than the later P3 components indexing higher-order context updating.

N-back results showed reduced P450 amplitude in MCI– relative to SCD in the 1-back condition, and absence of the PNwm component in MCI+, replicating prior findings [19, 20, 36] and indicating early disruption of working memory networks. Behaviourally, MCI+ showed elevated error rates and slower reaction times, while MCI– was indistinguishable from SCD, suggesting that electrophysiological markers (P450, PNwm) are detectable before behavioural impairment emerges.

### 4.2. ASSR and gamma synchrony: a non-monotonic trajectory

MCI– participants exhibited higher gamma ITC at Fz and Cz than both SCD and MCI+. This was unexpected given literature reporting higher ASSR gamma activity in AD [23, 24]; we had anticipated MCI+ to show the highest ITC. Instead, MCI– showed the highest values, with MCI+ comparable to SCD.

This non-monotonic trajectory may be interpreted within the framework of Gallego-Rudolf et al. [41], who describe how amyloid deposition initially accelerates neural activity before tau-driven degeneration leads to slowing. Heightened gamma ITC in ATN-negative MCI may thus reflect transient cortical hyperexcitability before tau pathology degrades network integrity — consistent with Sohrabpour et al. [28], who emphasize that EEG signatures of AD evolve in a non-monotonic fashion not aligned with progressive *Aβ* and tau accumulation. Future longitudinal studies across ATN-stratified cohorts will be essential to determine whether elevated gamma synchrony in MCI– represents a compensatory mechanism or a pre-existing trait.

### 4.3. Resting-state EEG features and MCI status characterisation

Resting-state analyses revealed that both MCI+ and SCD exhibited increased lower-frequency and decreased beta power compared to MCI–. This non-monotonic profile — with MCI– occupying an opposite position relative to both MCI+ and SCD — echoes Sohrabpour et al. [28], who note that spectral power inconsistencies across the literature likely reflect stage-dependent, non-linear dynamics rather than methodological noise.

PAF showed robust group differences, particularly in the eyes-closed condition, where MCI+ had reduced PAF compared to MCI– across multiple regions. MCI–, by contrast, showed higher PAF than SCD in parietal and occipital regions, placing it at the fast end of the three groups rather than on a monotonic slowing gradient. Our correlations confirmed that lower PAF associates with elevated tau, linking intrinsic cortical rhythm slowing to tau pathology specifically [15].

PLV analyses revealed frequency-specific network alterations: MCI+ showed reduced delta PLV between left temporal and occipital regions, indicating stage-specific low-frequency disconnection; both MCI– and MCI+ showed reduced beta PLV relative to SCD, reflecting broader network dysfunction preceding ATN positivity. EEG complexity was higher in MCI– than SCD across frontal, parietal, and occipital regions, consistent with the broader hyperexcitability profile.

### 4.4. EEG hyperexcitability in MCI–

Results from the ASSR, resting-state spectral, and complexity analyses converge on a consistent picture: MCI– participants display increased high-frequency synchrony, reduced low-frequency power, and higher signal complexity — consistent with cortical hyperexcitability or compensatory hypersynchrony in ATN-negative MCI.

This aligns with models by Maestú et al. [42] and Pusil et al. [43] suggesting early amyloid deposition promotes hypersynchronization, and mirrors Gaubert et al. [16], who showed early neurodegeneration is characterized by a “speeding up” of EEG rhythms interpreted as compensatory. Gallego-Rudolf et al. [41] further articulate that under amyloid influence, neural activity is initially accelerated; only as tau accumulates does activity slow and symptoms intensify. The MCI– group — cognitively impaired without ATN positivity — may represent this transitional state. This underscores the importance of biomarker-defined stratification: pooling MCI+ and MCI– would have obscured these opposing physiological profiles.

### 4.5. Resting-state oscillatory activity reflects pathological measures of cognition and biomarkers

Theta and beta rhythms during eyes-open rest showed complementary biomarker associations: higher theta correlated with lower MMSE and higher tau; higher beta correlated with higher amyloid and lower tau, consistent with EEG slowing in MCI and AD [44, 45]. PAF also tracked tau pathology, consistent with evidence that alpha slowing reflects tau-mediated cortical disconnection [46]. Higher SFR and lower Higuchi complexity were associated with elevated tau and reduced amyloid, capturing complementary aspects of neural integrity consistent with the loss of physiological complexity characteristic of AD [47, 48].

### 4.6. EEG as a non-invasive surrogate for biomarker assessment

These findings support EEG as a non-invasive proxy for biomarker status. Features including P3a amplitude, theta/beta power, PAF, SFR, and complexity could inform amyloid and tau pathology without lumbar puncture or blood sampling [7, 46]. Individual EEG features achieved AUCs of 0.72–0.78 — insufficient for standalone diagnosis but a meaningful signal above chance [16]. The added value of EEG was highly dependent on the diagnostic contrast: when distinguishing ATN– from ATN+, plasma pTau217 performed near ceiling (AUC = 0.96–0.99), leaving little room for incremental EEG contribution [49]. By contrast, for SCD versus MCI discrimination, plasma biomarkers were essentially uninformative (AUC *≈* 0.32–0.33), yet EEG alone matched MMSE performance (AUC *≈* 0.72–0.75). This underscores that while plasma markers are superior for detecting pathology, EEG is more sensitive to the functional transition from preclinical to symptomatic stages [11].

### 4.7. Conclusion

This study demonstrates that a structured multi-task EEG battery captures neurophysiological signatures tracking both cognitive status and amyloid and tau pathology across ATN-defined prodromal AD stages. ERPs indexed attentional and working memory dysfunction tied to amyloid and tau burden; resting-state spectral and connectivity markers reflected oscillatory slowing; and ASSR and complexity measures revealed a paradoxical hyperexcitability profile in ATN-negative MCI consistent with a non-monotonic, stage-dependent model of neurophysiological change.

Classification analyses revealed that EEG is most clinically valuable as a first-pass screening instrument at the clinical entry point — where patients present with subjective or mild cognitive complaints but plasma pTau217 and pTau181 may remain functionally silent. EEG can identify individuals warranting escalation to further biomarker evaluation, reducing unnecessary invasive procedures. Replication in larger, longitudinally followed cohorts with full ATN characterisation across all participants will be essential to validate this triage logic and to resolve whether the hyperexcitability pattern in MCI– represents a transient compensatory state or a stable phenotype. The COGBAT battery represents a promising, cost-effective, and scalable candidate for integration into memory clinic workflows, offering the potential to streamline clinical decision-making in the era of disease-modifying therapies.

## Data Availability

All data produced in the present study are available upon reasonable request to the authors

## Acknowledgements

We acknowledge and thank the participants in this study and their families. We give special thanks to Laia Cañada for managing the patient recruitment, Xavier Morató and Merce Boada for enabling the study at ACE Alzheimer Center Barcelona and Karan Chugani and Victor Fernández for conducting the EEG recordings.

## Consent Statement

All participants provided written informed consent prior to participation. The study was approved by the Ethics Committee of the Universitat Autònoma de Barcelona (UAB; reference CEEAH 6252) and by the Ethics Committee of the University Hospital of Bellvitge (CEIm; reference ICPS021/123).

## Supporting Information

Additional supporting information may be found in the online version of this article. **Figure S1.** Auditory oddball — P3b and N2b ERPs. **Figure S2.** N-back — PNwm ERP. **Figure S3.** Resting-state EEG complexity features, eyes-open condition. **Figure S4.** Correlations between EEG features, clinical scores, and biomarkers. **Table S1.** Behavioural performance results. **Table S2.** Significant resting-state band-power and slow-to-fast ratio ANOVA results. **Table S3.** Significant resting-state peak alpha frequency ANOVA results. **Table S4.** Significant eyes-closed connectivity ANOVA results. **Table S5.** Significant EEG complexity ANOVA results.

