## Supplementary material for "EEG Biomarkers for ATN Classification in Early Alzheimer’s Disease"

**Braboszcz et al.**

This document contains supplementary figures and tables referenced in the main article.

### Supplementary Figures

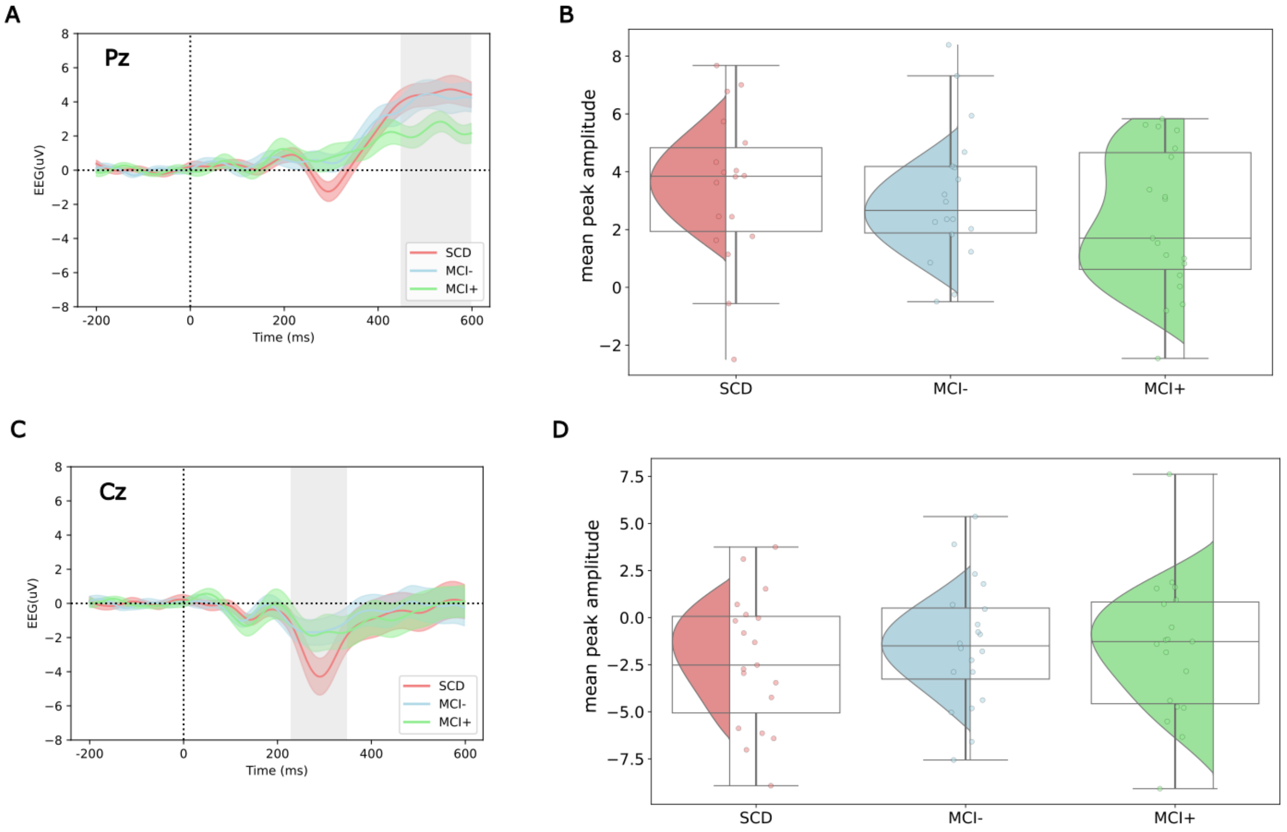


**Figure S1.** Auditory Oddball – P3b and N2b ERPs. **A.** P3b ERP on Pz for each group. The 450–600 ms time window of interest is shown as a grey area. **B.** Mean peak amplitude distribution of P3b on Pz for each group over the time window of interest. **C.** N2b ERP on Cz for each group. The 230–530 ms time window of interest is shown as a grey area. **D.** Mean peak amplitude distribution of N2b on Cz for each group over the time window of interest.

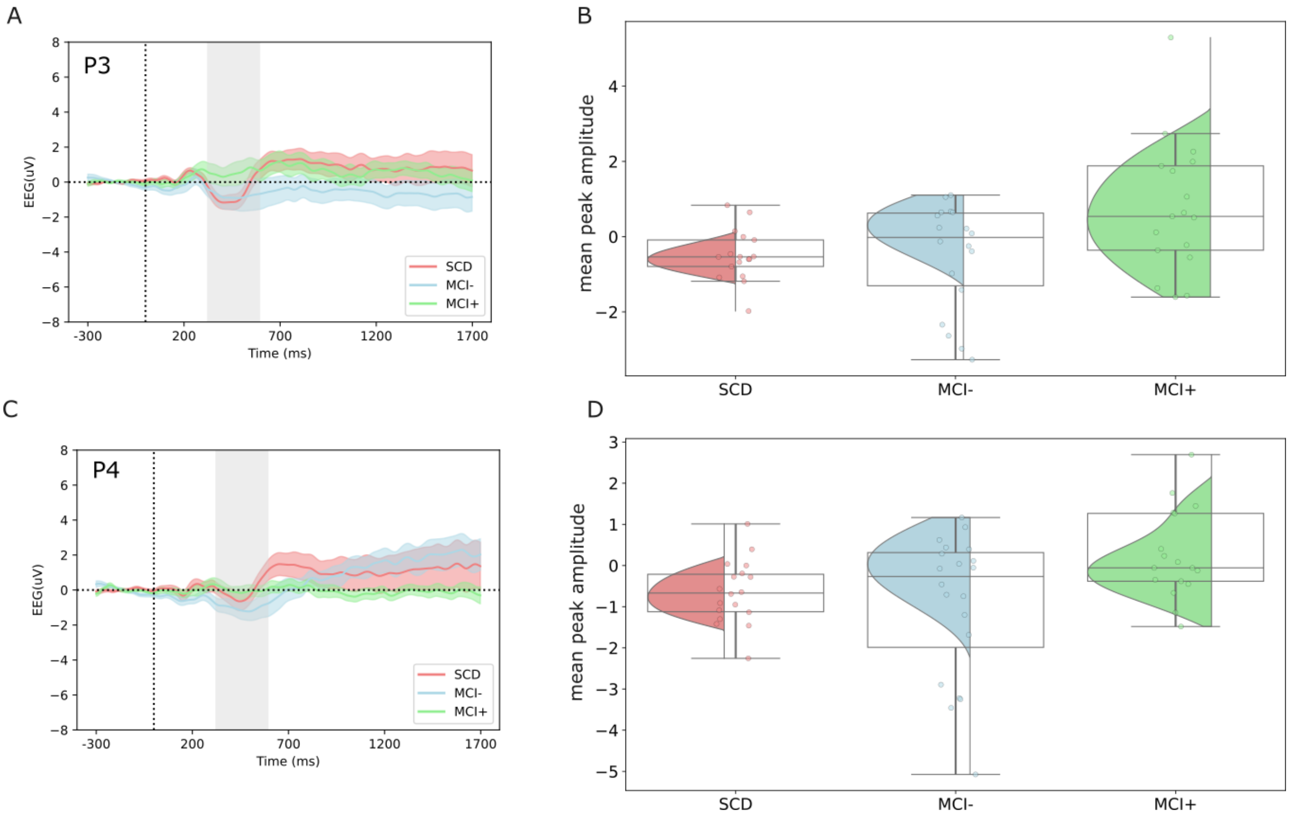


**Figure S2.** N-back – PNwm ERP. **A.** PNwm ERP on P3 for each group. The 320–597 ms time window of interest is shown as a grey area. **B.** Mean peak amplitude distribution of PNwm on P3 for each group over the time window of interest. The mean amplitude of the PNwm in MCI+ group is significantly reduced compared to SCD group ($p < 0.05$). **C.** PNwm ERP on P4 for each group. The 320–597 ms time window of interest is shown as a grey area. **D.** Mean peak amplitude distribution of PNwm on P4 for each group over the time window of interest. The mean amplitude of the PNwm in MCI+ group is significantly reduced compared to MCI$-$ group ($p < 0.05$).

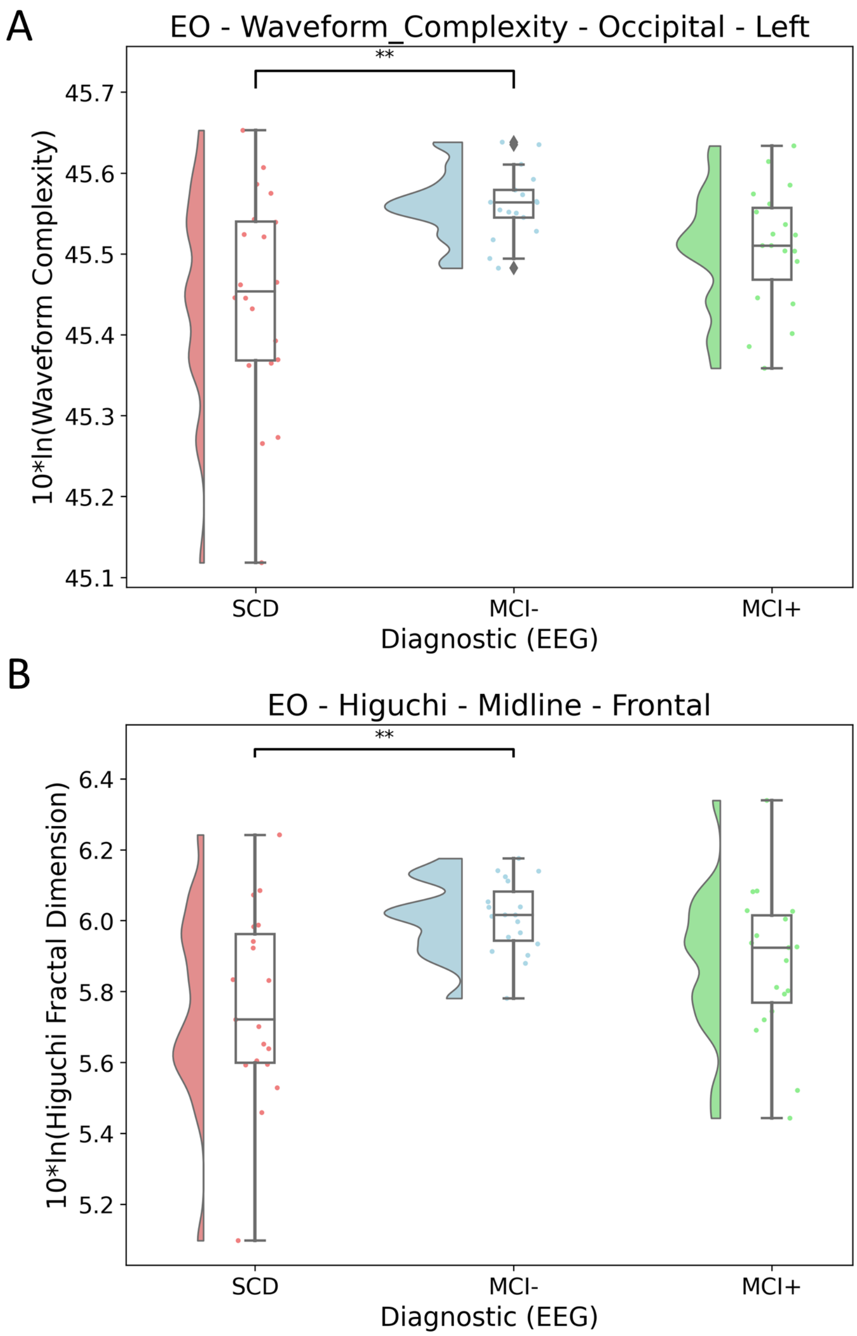


**Figure S3.** Resting-state EEG complexity features in the eyes-open condition. **A.** Barplot of Waveform Complexity at left occipital regions, showing higher complexity in MCI− compared to SCD. **B.** Barplot of Higuchi Fractal Dimension at midline frontal regions, indicating increased complexity in MCI− relative to SCD. In both panels, SCD is shown in red, MCI− in blue, and MCI+ in green. Asterisks indicate significance levels: * $p < 0.05$, ** $p < 0.01$, *** $p < 0.001$.

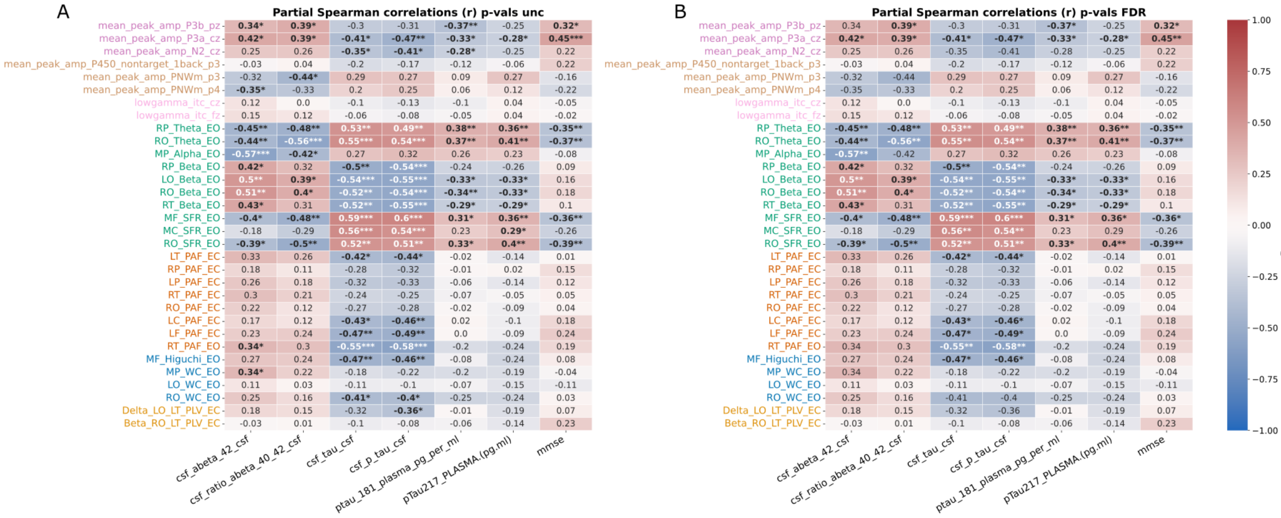


**Figure S4.** Correlation between EEG features and clinical scores and biomarkers. **A.** Correlation results with $p$-values uncorrected. **B.** Correlation results with $p$-values corrected for multiple comparisons using FDR. In both panels, partial Spearman correlations between EEG features and CSF/plasma biomarkers and MMSE scores are shown. The colour scale and numerical values show the Spearman $r$ value; statistical significance is indicated as: * $p < 0.05$, ** $p < 0.01$, *** $p < 0.001$.

### Supplementary Tables

**Table S1.** Behavioural performance results. Auditory Oddball accuracy was analysed with Welch's ANOVA and Games-Howell post-hoc tests. N-back accuracy and reaction time were analysed with mixed ANOVAs (Group × Condition). Effect sizes: $\eta_p^2$ (omnibus), Hedges' $g$ (contrasts).

| Task | Measure | $F(df_1, df_2)$ | $p_{unc}$ | $\eta_p^2$ | Contrast | $\Delta M$ | $p_{adj}$ | $g$ |
|---|---|---|---|---|---|---|---|---|
| **Auditory Oddball** | Accuracy (%) | 9.37 (2, 28.80) | < .001 | .320 | MCI− vs MCI+ | 20.41 | .010** | 1.023 |
| | | | | | MCI− vs SCD | −4.44 | .195 | −0.580 |
| | | | | | MCI+ vs SCD | −24.85 | .001** | −1.282 |
| **N-back** | *Accuracy (%) — Mixed ANOVA: Group × Condition* | | | | | | | |
| | Omnibus | 13.39 (2, 48)[a] | < .001 | .358 | Group (between) | — | — | — |
| | | 59.56 (1, 48)[b] | < .001 | .554 | Condition (within) | — | — | — |
| | | 6.28 (2, 48)[c] | .004 | .207 | Group × Condition | — | — | — |
| | Group | — | — | — | MCI+ vs MCI− | −7.83 | .008** | −1.140 |
| | | | | | MCI+ vs SCD | −9.35 | .004** | −1.485 |
| | | | | | MCI− vs SCD | −1.52 | .199 | −0.445 |
| | Group × Cond. | — | — | — | *0-back:* | | | |
| | | | | | MCI+ vs MCI− | −3.39 | .079 | −0.708 |
| | | | | | MCI+ vs SCD | −5.44 | .021* | −1.114 |
| | | | | | MCI− vs SCD | −2.05 | .074 | −0.712 |
| | | | | | *1-back:* | | | |
| | | | | | MCI+ vs MCI− | −12.33 | .014* | −1.156 |
| | | | | | MCI+ vs SCD | −13.53 | .012* | −1.382 |
| | | | | | MCI− vs SCD | −1.20 | .456 | −0.252 |
| | *Reaction Time (sec) — Mixed ANOVA: Group × Condition* | | | | | | | |
| | Omnibus | 13.44 (2, 51)[d] | < .001 | .345 | Group (between) | — | — | — |
| | | 49.27 (1, 51)[e] | < .001 | .491 | Condition (within) | — | — | — |
| | | 3.23 (2, 51)[f] | .048 | .112 | Group × Condition | — | — | — |
| | Group | — | — | — | MCI+ vs MCI− | 0.096 | < .001*** | 1.637 |
| | | | | | MCI+ vs SCD | 0.086 | .001** | 1.195 |
| | | | | | MCI− vs SCD | −0.010 | .306 | −0.325 |
| | Group × Cond. | — | — | — | *0-back:* | | | |
| | | | | | MCI+ vs MCI− | 0.067 | .005** | 1.059 |
| | | | | | MCI+ vs SCD | 0.086 | .005** | 1.078 |
| | | | | | MCI− vs SCD | 0.018 | .977 | 0.009 |
| | | | | | *1-back:* | | | |
| | | | | | MCI+ vs MCI− | 0.125 | < .001*** | 1.990 |
| | | | | | MCI+ vs SCD | 0.088 | .003** | 1.164 |
| | | | | | MCI− vs SCD | −0.037 | .071 | −0.606 |

[*] $p < .05$; [**] $p < .01$; [***] $p < .001$.
$\Delta M$ = mean difference $g$ = Hedges' $g$; $\eta_p^2$ = partial eta-squared; Cond. = Condition; — = not applicable.

**Table S2.** Significant resting-state ANOVA results and post-hoc Tukey HSD for relative band power and slow-to-fast ratio. Effect sizes: $\eta_p^2$ (omnibus), Hedges' $g$ (contrasts). $\Delta M$ is the mean difference of the first minus the second group of each contrast. * $p < .05$; ** $p < .01$; *** $p < .001$.

| Band | ROI | F(df1,df2) | $p_{FDR}$ | $\eta_p^2$ | Contrast | $\Delta M$ | $p_{adj}$ | g |
|---|---|---|---|---|---|---|---|---|
| Theta | Parietal Right | 5.77 (2,53) | 0.04* | 0.18 | MCI− vs MCI+ | −3.358 | 0.005** | −1.265 |
| | | | | | SCD vs MCI+ | −2.427 | 0.047* | −0.718 |
| | | | | | SCD vs MCI− | 0.930 | 0.622 | 0.282 |
| Theta | Occipital Right | 5.98 (2,55) | 0.04* | 0.18 | MCI− vs MCI+ | −3.271 | 0.005** | −1.040 |
| | | | | | SCD vs MCI+ | −2.647 | 0.030* | −0.803 |
| | | | | | SCD vs MCI− | 0.624 | 0.807 | 0.201 |
| Theta | Midline Frontal | 5.32 (2,52) | 0.04* | 0.17 | MCI− vs MCI+ | −3.538 | 0.006** | −1.400 |
| | | | | | SCD vs MCI+ | −1.313 | 0.430 | −0.354 |
| | | | | | SCD vs MCI− | 2.225 | 0.103 | 0.634 |
| Theta | Midline Central | 6.40 (2,53) | 0.04* | 0.19 | MCI− vs MCI+ | −3.397 | 0.003** | −1.245 |
| | | | | | SCD vs MCI+ | −0.779 | 0.703 | −0.238 |
| | | | | | SCD vs MCI− | 2.618 | 0.025* | 0.845 |
| Alpha | Midline Parietal | 5.42 (2,52) | 0.04* | 0.17 | MCI− vs MCI+ | −2.848 | 0.008** | −1.163 |
| | | | | | SCD vs MCI+ | −2.143 | 0.049* | −0.708 |
| | | | | | SCD vs MCI− | 0.704 | 0.721 | 0.248 |
| Beta | Temporal Right | 6.20 (2,55) | 0.04* | 0.18 | MCI− vs MCI+ | 3.772 | 0.003** | 1.201 |
| | | | | | SCD vs MCI+ | 2.051 | 0.144 | 0.542 |
| | | | | | SCD vs MCI− | −1.721 | 0.233 | −0.547 |
| Beta | Parietal Right | 6.06 (2,55) | 0.04* | 0.18 | MCI− vs MCI+ | 3.981 | 0.003** | 1.187 |
| | | | | | SCD vs MCI+ | 1.862 | 0.244 | 0.491 |
| | | | | | SCD vs MCI− | −2.119 | 0.148 | −0.582 |
| Beta | Occipital Left | 5.87 (2,55) | 0.04* | 0.18 | MCI− vs MCI+ | 3.724 | 0.005** | 1.321 |
| | | | | | SCD vs MCI+ | 1.027 | 0.639 | 0.260 |
| | | | | | SCD vs MCI− | −2.697 | 0.046* | −0.711 |
| Beta | Occipital Right | 7.40 (2,56) | 0.04* | 0.21 | MCI− vs MCI+ | 4.161 | 0.001** | 1.436 |
| | | | | | SCD vs MCI+ | 1.124 | 0.580 | 0.279 |
| | | | | | SCD vs MCI− | −3.037 | 0.022* | −0.818 |
| Beta | Midline Frontal | 13.11 (2,53) | 0.002** | 0.33 | MCI− vs MCI+ | 3.568 | 0.002** | 1.231 |
| | | | | | SCD vs MCI+ | −1.418 | 0.359 | −0.402 |
| | | | | | SCD vs MCI− | −4.986 | <.001*** | −1.659 |
| Beta | Midline Central | 7.49 (2,55) | 0.04* | 0.21 | MCI− vs MCI+ | 3.696 | 0.003** | 1.171 |
| | | | | | SCD vs MCI+ | 0.405 | 0.924 | 0.108 |
| | | | | | SCD vs MCI− | −3.291 | 0.008** | −1.048 |
| Beta | Midline Parietal | 5.30 (2,52) | 0.04* | 0.17 | MCI− vs MCI+ | 3.273 | 0.013* | 1.117 |
| | | | | | SCD vs MCI+ | 0.366 | 0.942 | 0.110 |
| | | | | | SCD vs MCI− | −2.907 | 0.025* | −0.757 |
| SFR | Occipital Right | 6.25 (2,53) | 0.04* | 0.19 | MCI− vs MCI+ | −4.500 | 0.003** | −1.030 |
| | | | | | SCD vs MCI+ | −3.239 | 0.054 | −0.756 |
| | | | | | SCD vs MCI− | 1.261 | 0.619 | 0.332 |
| SFR | Midline Frontal | 6.94 (2,55) | 0.04* | 0.20 | MCI− vs MCI+ | −5.442 | 0.003** | −1.224 |
| | | | | | SCD vs MCI+ | −1.076 | 0.774 | −0.201 |
| | | | | | SCD vs MCI− | 4.366 | 0.018* | 0.872 |
| SFR | Midline Central | 5.59 (2,54) | 0.04* | 0.17 | MCI− vs MCI+ | −5.011 | 0.012* | −1.028 |
| | | | | | SCD vs MCI+ | −0.252 | 0.988 | −0.045 |
| | | | | | SCD vs MCI− | 4.759 | 0.018* | 0.884 |

**Table S3.** Significant resting-state ANOVA results and post-hoc Tukey HSD tests for peak alpha frequency after FDR correction. Effect sizes: $\eta_p^2$ (omnibus), Hedges' $g$ (contrasts). * $p < .05$; ** $p < .01$; *** $p < .001$.

| Cond. | ROI | $F(df_1, df_2)$ | $p_{FDR}$ | $\eta_p^2$ | Contrast | $\Delta M$ | $p_{adj}$ | $g$ |
|---|---|---|---|---|---|---|---|---|
| EO | Temporal Right | 5.36 (2,55) | 0.049* | 0.16 | MCI− vs MCI+ | 1.662 | <.001*** | 1.228 |
| | | | | | SCD vs MCI+ | 1.032 | 0.039* | 0.830 |
| | | | | | SCD vs MCI− | −0.630 | 0.264 | −0.496 |
| EC | Frontal Left | 6.54 (2,51) | 0.010** | 0.20 | MCI− vs MCI+ | 0.980 | 0.003** | 1.038 |
| | | | | | SCD vs MCI+ | 0.582 | 0.129 | 0.753 |
| | | | | | SCD vs MCI− | −0.397 | 0.337 | −0.459 |
| EC | Temporal Right | 4.44 (2,56) | 0.023* | 0.14 | MCI− vs MCI+ | 0.930 | 0.018* | 0.906 |
| | | | | | SCD vs MCI+ | 0.224 | 0.762 | 0.199 |
| | | | | | SCD vs MCI− | −0.707 | 0.082 | −0.880 |
| EC | Temporal Left | 5.87 (2,52) | 0.011* | 0.18 | MCI− vs MCI+ | 1.044 | 0.004** | 0.998 |
| | | | | | SCD vs MCI+ | 0.566 | 0.177 | 0.565 |
| | | | | | SCD vs MCI− | −0.478 | 0.258 | −0.579 |
| EC | Parietal Right | 7.17 (2,50) | 0.010** | 0.22 | MCI− vs MCI+ | 0.915 | 0.002** | 1.152 |
| | | | | | SCD vs MCI+ | 0.007 | 0.999 | 0.009 |
| | | | | | SCD vs MCI− | −0.908 | 0.002** | −1.524 |
| EC | Parietal Left | 8.63 (2,49) | 0.008** | 0.26 | MCI− vs MCI+ | 1.026 | 0.005** | 1.430 |
| | | | | | SCD vs MCI+ | 0.131 | 0.902 | 0.124 |
| | | | | | SCD vs MCI− | −0.895 | 0.013* | −0.930 |
| EC | Occipital Right | 5.82 (2,55) | 0.011* | 0.17 | MCI− vs MCI+ | 0.589 | 0.069 | 0.710 |
| | | | | | SCD vs MCI+ | −0.104 | 0.916 | −0.135 |
| | | | | | SCD vs MCI− | −0.693 | 0.025* | −0.785 |
| EC | Central Left | 6.82 (2,51) | 0.010** | 0.21 | MCI− vs MCI+ | 0.962 | 0.005** | 1.008 |
| | | | | | SCD vs MCI+ | 0.631 | 0.089 | 0.668 |
| | | | | | SCD vs MCI− | −0.332 | 0.489 | −0.401 |

**Table S4.** Significant eyes-closed connectivity ANOVA results and post-hoc Tukey HSD tests after FDR correction. Effect sizes: $\eta_p^2$ (omnibus), Hedges' $g$ (contrasts). * $p < .05$; ** $p < .01$; *** $p < .001$.

| Band | Connection | $F(df_1, df_2)$ | $p_{FDR}$ | $\eta_p^2$ | Contrast | $\Delta M$ | $p_{adj}$ | $g$ |
|---|---|---|---|---|---|---|---|---|
| Delta | Temporal L – Occipital L | 9.99 (2,54) | 0.044* | 0.27 | MCI− vs MCI+ | 0.067 | <.001*** | 1.305 |
| | | | | | SCD vs MCI+ | 0.055 | 0.003** | 1.311 |
| | | | | | SCD vs MCI− | −0.012 | 0.708 | −0.226 |
| Beta | Occipital R – Temporal L | 12.70 (2,49) | 0.016* | 0.34 | MCI− vs MCI+ | −0.014 | 0.706 | −0.390 |
| | | | | | SCD vs MCI+ | 0.063 | 0.002** | 1.182 |
| | | | | | SCD vs MCI− | 0.077 | <.001*** | 1.312 |

**Table S5.** Significant EEG complexity ANOVA results and post-hoc Tukey HSD tests after FDR correction. Effect sizes: $\eta_p^2$ (omnibus), Hedges' $g$ (contrasts). * $p < .05$; ** $p < .01$; *** $p < .001$.

| **Measure** | **Region** | $F(df_1, df_2)$ | $p_{FDR}$ | $\eta_p^2$ | **Contrast** | $\Delta M$ | $p_{adj}$ | $g$ |
|---|---|---|---|---|---|---|---|---|
| Higuchi | Frontal Midline | 6.922 (2,54) | 0.027* | 0.204 | MCI− vs MCI+ | 0.130 | 0.134 | 0.774 |
| | | | | | SCD vs MCI+ | −0.118 | 0.191 | −0.478 |
| | | | | | SCD vs MCI− | −0.248 | 0.001** | −1.191 |
| Waveform | Occipital Left | 6.405 (2,54) | 0.021* | 0.192 | MCI− vs MCI+ | 0.047 | 0.283 | 0.721 |
| | | | | | SCD vs MCI+ | −0.061 | 0.113 | −0.553 |
| | | | | | SCD vs MCI− | −0.108 | 0.002** | −1.044 |
| | Occipital Right | 5.818 (2,53) | 0.023* | 0.180 | MCI− vs MCI+ | 0.072 | 0.118 | 0.938 |
| | | | | | SCD vs MCI+ | −0.046 | 0.386 | −0.371 |
| | | | | | SCD vs MCI− | −0.118 | 0.004** | −0.990 |
| | Midline Parietal | 7.589 (2,54) | 0.016* | 0.219 | MCI− vs MCI+ | 0.058 | 0.178 | 0.733 |
| | | | | | SCD vs MCI+ | −0.065 | 0.102 | −0.577 |
| | | | | | SCD vs MCI− | −0.123 | 0.001** | −1.196 |
